# A Minimal PBPK Model Describes the Differential Disposition of Silica Nanoparticles In Vivo

**DOI:** 10.1101/2024.09.18.24313941

**Authors:** Madison Parrot, Joseph Cave, Maria J Pelaez, Hamidreza Ghandehari, Prashant Dogra, Venkata Yellepeddi

## Abstract

Nanoparticles (NPs) have emerged as promising candidates for drug delivery due to their tunable physical and chemical properties. Among these, silica nanoparticles (SiNPs) are particularly valued for their biocompatibility and adaptability in applications like drug delivery and medical imaging. However, predicting SiNP biodistribution and clearance remains a significant challenge. To address this, we developed a minimal physiologically-based pharmacokinetic (mPBPK) model to simulate the systemic disposition of SiNPs, calibrated using in vivo PK data from mice. The model assesses how variations in surface charge, size, porosity, and geometry influence SiNP biodistribution across key organs, including the kidneys, lungs, liver, and spleen. A global sensitivity analysis identified the most influential parameters, with the unbound fraction and elimination rate constants for the kidneys and MPS emerging as critical determinants of SiNP clearance. Non-compartmental analysis (NCA) further revealed that aminated SiNPs exhibit high accumulation in the liver, spleen, and kidneys, while mesoporous SiNPs primarily accumulate in the lungs. Rod-shaped SiNPs showed faster clearance compared to spherical NPs. The mPBPK model was extrapolated to predict SiNP behavior in humans, yielding strong predictive accuracy with Pearson correlation coefficients of 0.98 for mice and 0.92 for humans. This model provides a robust framework for predicting the pharmacokinetics of diverse SiNPs, offering valuable insights for optimizing NP-based drug delivery systems and guiding the translation of these therapies from preclinical models to human applications.

## Introduction

In recent years, SiNPs have garnered significant attention due to their unique physical and chemical properties, making them valuable in drug delivery applications.(1) Their large surface area to volume ratio allows facile surface modification and conjugation, enhancing their utility in targeting and delivering therapeutic agents.(2) For instance, functionalized SiNPs have demonstrated remarkable potential in imaging and targeted delivery to improve therapeutic efficacy and minimize side effects due to their tunable characteristics and biocompatibility.(3-6) Despite their versatility, a significant unmet need remains in translating these preclinical outcomes into clinical practice, as understanding their behavior in complex biological systems and addressing interspecies differences continue to pose challenges.

SiNPs comprise silicon dioxide (SiO_2_), a compound where silicon atoms are covalently bonded to oxygen atoms, forming a robust tetrahedral network and three-dimensional lattice structure.(7) This network of SiO_2_ can self-assemble into various nanostructures, including NPs with unique properties depending on their size, shape, porosity, and surface modifications.(7) Among the various types of SiNPs, mesoporous SiNPs have potential as drug-delivery vehicles and diagnostics due to their adjustable pore size, tunable size, and geometry.(8) SiNPs can be included in gold or iron oxide composites for plasmonic and thermal ablation therapy.(9, 10) Biomedical applications of SiNPs have been extensively reviewed elsewhere.(11, 12) Significant applications of SiNPs include drug delivery, diagnostics, and imaging.

Pharmacokinetics (PK) studies how compounds are absorbed, distributed, metabolized, and excreted in biological systems. Knowledge of NP PK is essential for developing safe and effective NP-based therapies. Unlike traditional small molecules, NPs exhibit complex behaviors influenced by their size, geometry, surface properties, and unique interactions with biological components.(13-15) NPs are known to accumulate in the liver or spleen due to uptake by the resident macrophages in the sinusoidal blood vessels.(16) Larger NPs, or those with a positive surface charge, show the highest uptake by the mononuclear phagocytic system (MPS).(17) Kumar et al. reviewed NP data from the literature to establish disposition trends and determined that SiNPs show higher lung, lower spleen, and lower heart concentrations than other NP types.(18)

The complexities of SiNPs necessitate advanced modeling approaches to predict NP PK accurately. PK modeling is a vital tool to understand NP behavior *in vivo*.(19, 20) The translation of NPs has only seen moderate success, highlighting the need for innovative methods in experimental design.(21-23) It has been established that NP characteristics do not consistently correlate with specific PK properties, particularly across NP types.(24) This necessitates using a PK modeling strategy for each NP type to establish the relationship between their physicochemical properties and PK parameters.

Physiologically-based pharmacokinetic (PBPK) models offer a sophisticated approach to understanding PK of therapeutics by incorporating detailed physiological and biochemical data.(25, 26) A whole-body PBPK model comprises several organ compartments where the compartments are connected via blood and lymph flow rates, with the transport phenomena mathematically characterized by a set of ordinary differential equations (ODEs). The ODEs contain known physiological parameters for the species of interest and unknown empirical and/or mechanistic parameters that are fitted using the observed data. These models simulate the whole-body behavior of therapeutics, providing insights into their PK profiles. PBPK models are widely utilized in drug development to bridge the gap between preclinical and clinical phases.(27) PBPK models find utility for optimizing the first-in-human dose to support the design of clinical trials.(28)

PBPK models are particularly valuable for NPs, allowing researchers to account for their unique properties and biological interactions within the body. Despite the advancements in PBPK modeling, there remains a gap in the literature regarding models tailored explicitly for SiNPs. To our knowledge, no PBPK models of SiNPs in mice or humans are reported in the literature. We thus developed a PBPK model to evaluate SiNPs of different sizes, surface charges, and geometries for the first time. The primary objective of this study is to develop a minimal PBPK (mPBPK) model for six types of SiNPs in mice and extrapolate the animal model to humans to describe NP-specific characteristics that affect NP biodistribution.

## Methods

### Preclinical and Clinical NP PK Data

The six NP types administered in mice are either nonporous silica nanospheres (Stöber) or mesoporous SiO_2_. The six NP types are mesoporous nanospheres (Meso), aminated mesoporous nanospheres (MA), Stöber nanospheres (Stöber), aminated Stöber nanospheres (SA), mesoporous nanorods with an aspect ratio of 8 (AR8), and aminated mesoporous nanorods with an aspect ratio of 8 (8A). These NPs were synthesized according to previously reported methods.(29) All NPs were radiolabeled with ^125^I according to methods described previously.(30) We extracted *in vivo* PK data for six different SiNPs were extracted using PlotDigitizer 2.6.9 9 (https://sourceforge.net/projects/plotdigitizer/files/plotdigitizer/).

The physicochemical characteristics of SiNPs investigated in this study are described in Table 1. A previous publication can find information on the synthesis, complete characterization, surface modifications, and subsequent labeling and biodistribution data of the six SiNPs used in mice.(30) Briefly, mice were injected with 20 mg/kg of NP suspension through the lateral tail vein for the PK studies. Blood and organs were harvested at 5 mins, 30 mins, 2, 24, and 72 hours. Radioactivity of the harvested organs was measured using a gamma counter and reported as the percent of injected dose per gram of tissue (%ID/g). Urine and feces were collected at 2, 24, 48, and 72 hours. Radioactivity from urine and feces was reported as the percentage of injected dose (%ID). The stability of the radioligand on SiNPs in mouse serum was confirmed for up to 72 hours using thin layer chromatography (TLC).

**Table 1.** Nanoparticle characterization. The NPs evaluated are mesoporous nanospheres (Meso), aminated mesoporous nanospheres (MA), Stöber nanospheres (Stöber), aminated Stöber nanospheres (SA), mesoporous nanorods with an aspect ratio of 8 (AR8), aminated mesoporous nanorods with an aspect ratio of 8 (8A), and Cornell dots (C dots). Surface charge is ranked as highly negative (−60 to −40 mV; symbol ---), moderately negative (−40 to −30 mV; symbol --), moderately positive (+10 to +20 mV; symbol ++), highly positive (+20 to +40 mV; symbol +++). N/A indicates not available.

| Nanoparticle | NP size by TEM*<br>(nm) | Surface Charge | Surface Modification | Porosity | Shape |
| --- | --- | --- | --- | --- | --- |
| Meso | 120 | -- | None | Mesoporous | Sphere |
| MA | 120 | +++ | Amine | Mesoporous | Sphere |
| Stöber | 115 | --- | None | Nonporous | Sphere |
| SA | 115 | ++ | Amine | Nonporous | Sphere |
| AR8 | 136 x 1028 | -- | None | Mesoporous | Rod |
| 8A | 136 x 1028 | +++ | Amine | Mesoporous | Rod |
| C dots | 6 | N/A | cRGDY peptides,<br>PEG,<br>Cy5 | Nonporous | Sphere |

\*TEM- transmission electron microscopy

The Cornell dots (^124^I-cRGDY–PEG–C dots) are Stöber SiNPs modified with a peptide chain, polyethylene glycol (PEG), and ^124^I for positron emission tomography (PET) imaging.(31) Full characterization of Cornell dots (C dots) can be found in Phillips et al.(31) Axial PET imaging for detection of the C dots was completed at 3, 24, and 72 hours after intravenous administration of 185 mega becquerels (MBq) (∼3.4 to 6.7 nmol) for each human subject (n=5). Radioactivity was reported as %ID/g.(31) The size of the cRGDY–PEG–Cy5 coatings were between 44 and 158 kDa.(32) The contribution to the overall weight of the radioactive tag to C dots was considered to be negligible.

### Model Development

We developed a minimal PBPK (mPBPK) model to simulate the whole-body biodistribution and clearance of SiNPs following intravenous injection. By emphasizing the biodistribution and clearance of SiNPs, which are directly influenced by their physicochemical properties, the model provides insights into how NP design affects PK. The model includes seven key compartments: plasma, the mononuclear phagocytic system (MPS, comprising the liver and spleen), lungs, kidneys, urine, feces, and the rest of the body (‘others’). A visual representation of the model can be seen in Figure 1.

**Figure 1.**
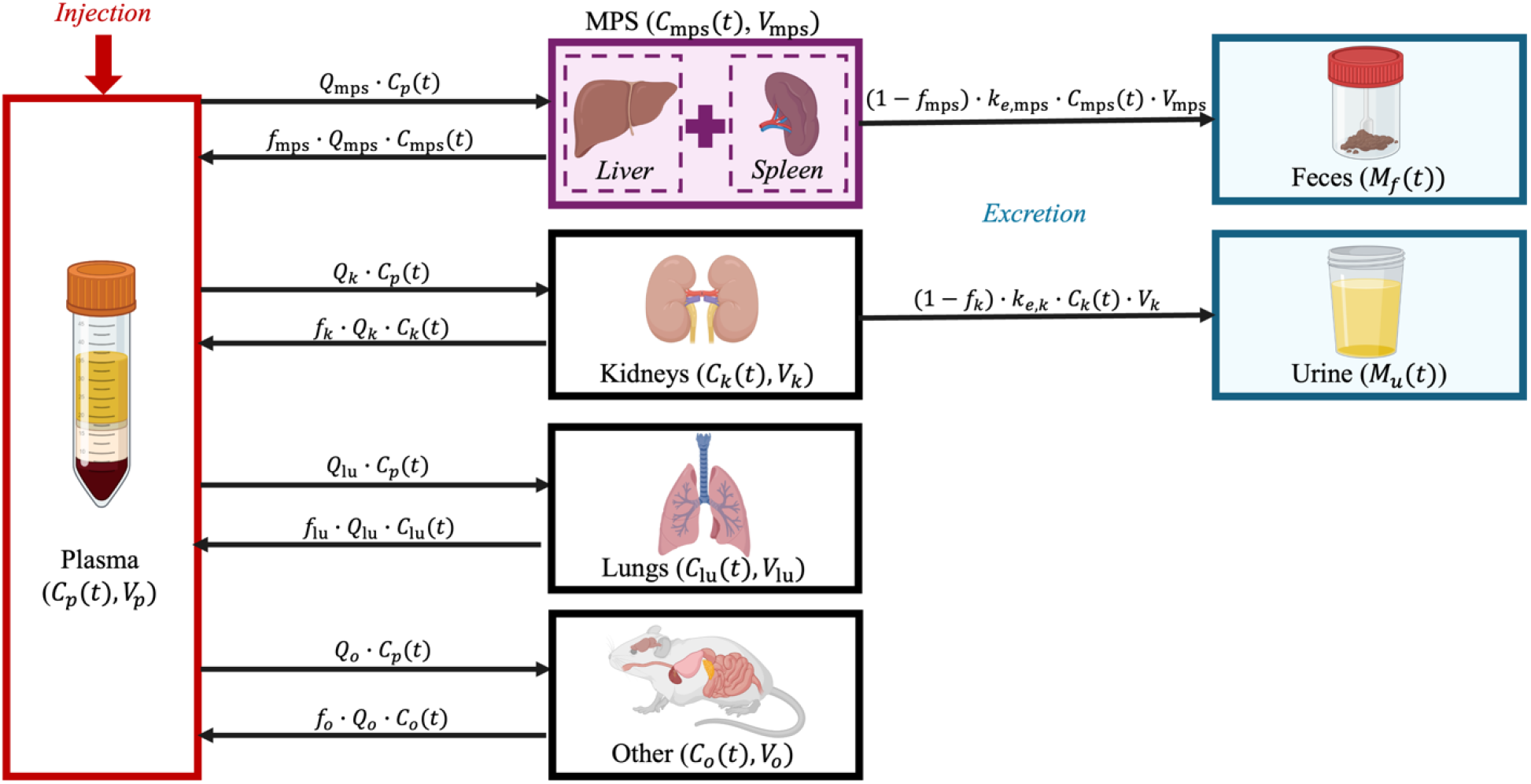
Illustration of the minimal PBPK (mPBPK) model to study the biodistribution and clearance of SiNPs across various compartments. This model quantifies SiNP concentrations in plasma (*C*_*p*_(*t*)), the MPS (*C*_mps_(*t*)), lungs (*C*_lu_(*t*)), kidneys (*C*_*k*_(*t*)), and other tissues (*C*_*o*_(*t*)). Each compartment, indexed by *i*, is characterized by its blood flow rate (*Q*_*i*_), the fraction of unbound nanoparticles (*f*_*i*_), and volume (*V*_*i*_). The model also details nanoparticle excretion through feces (*M*_*f*_(*t*)) and urine (*M*_*u*_(*t*)), with clearance rates from the MPS (*k*_*e*,mps_) and kidneys (*k*_*e,k*_), respectively.

A system of ordinary differential equations (ODEs, Equations 1-7) was used to describe the concentration kinetics of SiNPs following intravenous administration. The model accounts for both perfusion-limited transport and first-order excretion processes. As shown in Figure 1, SiNPs are distributed to different organs based on perfusion, which is determined by the blood flow rate *Q*_*i*_ for each organ *i*. The influx of SiNPs into each organ is proportional to the blood flow rate *Q*_*i*_ and the concentration of SiNPs in the plasma, *C*_*p*_(*t*). The outflux is proportional to the concentration of freely circulating (i.e., unbound) SiNPs in the organ, represented by *f*_*i*_ · *C*_*i*_(*t*), where *C*_*i*_(*t*) is the total concentration of SiNPs in organ *i* and *f*_*i*_ is the fraction of unbound SiNPs.This transport process is modeled as first-order, where both the influx and outflux are governed by perfusion-limited kinetics.

The excretion of SiNPs occurs through the MPS and kidneys, both of which follow first-order excretion kinetics. The excretion term depends on the concentration of bound (i.e., non-circulating) SiNPs in each organ, modulated by an organ-specific excretion rate constant,*k*_*e,i*,_. The fraction of bound SiNPs, 1− *f*_*i*_, is used to define the excretion rate as (1− *f*_*i*_) · *k*_*e,i*_ · *C*_*i*_(*t*). In the MPS, SiNPs are excreted into feces, while in the kidneys, they are excreted into urine.

The system of ODEs used to describe the concentration and mass kinetics of SiNPs in different compartments is as follows:

#### SiNP concentration kinetics in plasma

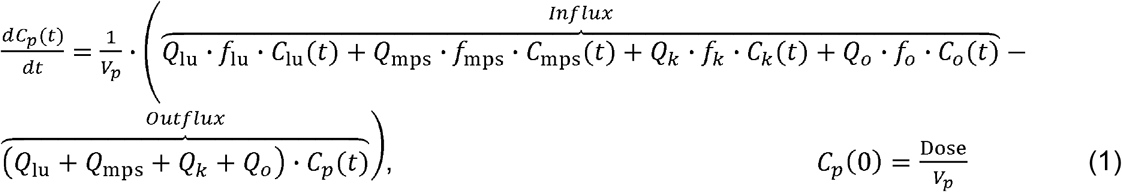

where, *C*_*p*_(*t*),*C*_lu_(*t*),*C*_mps_(*t*),*C*_*k*_(*t*), and *C*_*o*_(*t*) are the SiNP concentrations in plasma, lungs, MPS organs, kidneys, and other compartments, respectively; *V*_*p*_ is the volume of the plasma compartment; *Q*_lu_,*Q*_mps_,*Q*_*k*_, and *Q*_*o*_ represent the blood flow rates of lungs, MPS organs kidneys, and other compartments, respectively; *f*_lu_, *f*_mps_, *f*_*k*_, and *f*_*o*_ are the fractions of freely circulating or unbound SiNPs in lungs, MPS organs, kidneys, and other compartments, respectively; is the injected Dose of SiNPs.

#### SiNP concentration kinetics in MPS

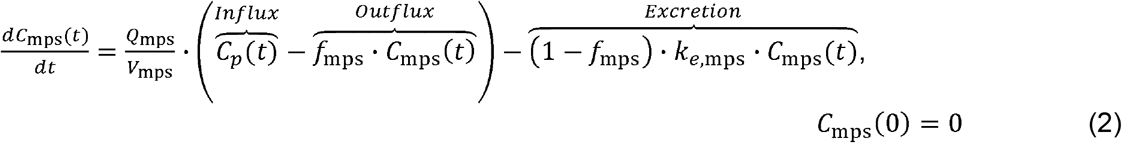

where, *V*_mps_ is the volume of the MPS compartment, *k*_*e*,mps_ is the excretion rate constant from the MPS into the feces.

#### SiNP concentration kinetics in lungs

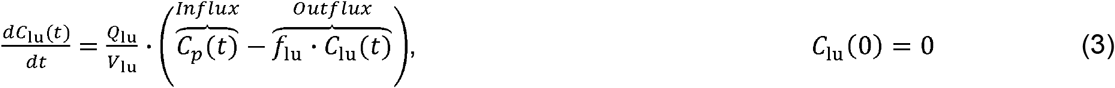

where, *V*_lu_ is the volume of the lungs compartment.

#### SiNP concentration kinetics in kidneys

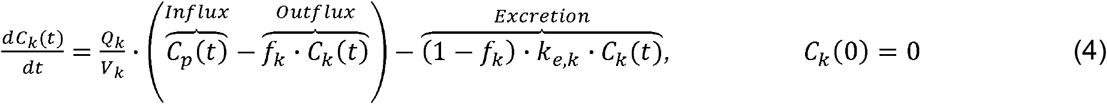

where, *V*_*k*_ is the volume of the kidneys compartment; *k*_*e,k*_ is the excretion rate constant from the kidneys into the urine.

#### SiNP concentration kinetics in others

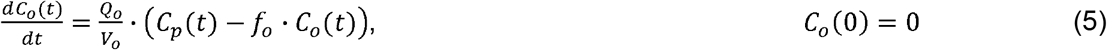

where, *V*_o_ is the volume of the others compartment.

#### SiNP mass kinetics in urine

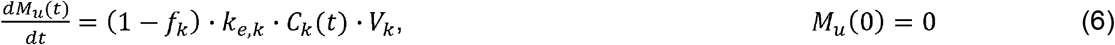

where, *M*_*u*_(*t*) represents the mass of SiNPs excreted into the urine.

#### SiNP mass kinetics in feces

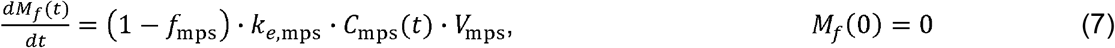

where, *M*_*f*_(*t*) represents the mass of SiNPs excreted into the feces.

### Numerical Solution and Parameter Estimation

The model was solved numerically using MATLAB (version R2024a, Natick, Massachusetts: The MathWorks Inc.) as an initial value problem. Initial conditions assume 100% of the injected dose in the plasma at time t=0, with zero concentration in the other compartments. The system of ODEs was solved using *ode15s* solver, suitable for stiff systems. Eight key parameters were fitted based on observed experimental data: the fraction unbound in the lungs (*f*_lu_), kidneys (*f*_*k*_), MPS organs (*f*_mps_), and other compartments (*f*_o_); the excretion rate constants from the kidneys (*k*_*e,k*_) and MPS organs (*k*_*e*,mps_); and the blood flow rates for the MPS (*Q*_mps_) and other compartments (*Q*_*o*_). These parameters were fitted using non-linear least squares fitting via the *lsqcurvefit* function in MATLAB. The fitting process was constrained within physiological bounds, ensuring that parameters were non-negative, and the fraction unbound did not exceed 1. The remaining physiological parameters were obtained directly from literature (Table 2).

**Table 2.**
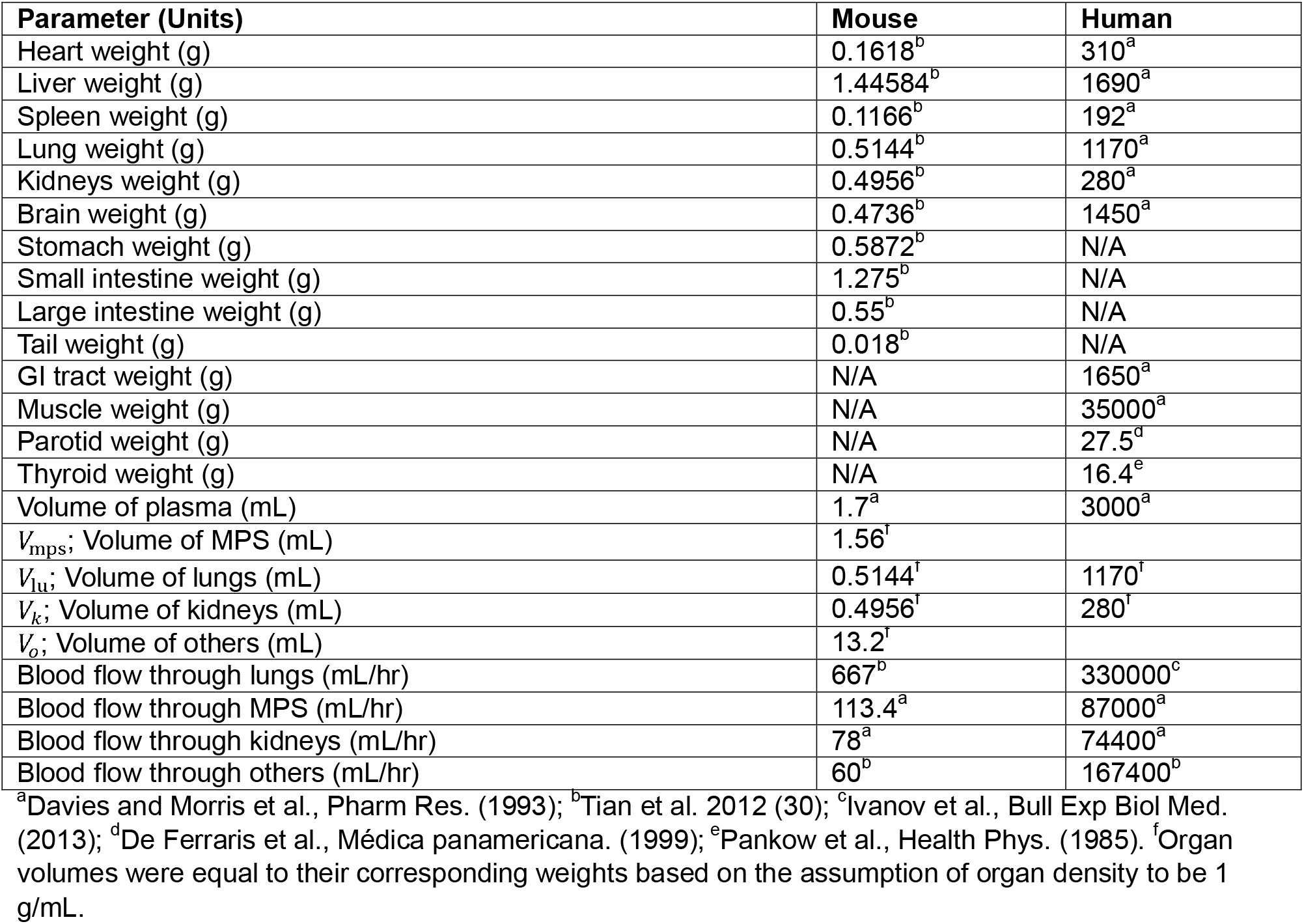
Physiological parameter values used in the PBPK model. N/A indicates that a parameter is not applicable to the respective model.

| Parameter (Units) | Mouse | Human |
| --- | --- | --- |
| Heart weight (g) | 0.1618<sup>b</sup> | 310<sup>a</sup> |
| Liver weight (g) | 1.44584<sup>b</sup> | 1690<sup>a</sup> |
| Spleen weight (g) | 0.1166<sup>b</sup> | 192<sup>a</sup> |
| Lung weight (g) | 0.5144<sup>b</sup> | 1170<sup>a</sup> |
| Kidneys weight (g) | 0.4956<sup>b</sup> | 280<sup>a</sup> |
| Brain weight (g) | 0.4736<sup>b</sup> | 1450<sup>a</sup> |
| Stomach weight (g) | 0.5872<sup>b</sup> | N/A |
| Small intestine weight (g) | 1.275<sup>b</sup> | N/A |
| Large intestine weight (g) | 0.55<sup>b</sup> | N/A |
| Tail weight (g) | 0.018<sup>b</sup> | N/A |
| GI tract weight (g) | N/A | 1650<sup>a</sup> |
| Muscle weight (g) | N/A | 35000<sup>a</sup> |
| Parotid weight (g) | N/A | 27.5<sup>d</sup> |
| Thyroid weight (g) | N/A | 16.4<sup>e</sup> |
| Volume of plasma (mL) | 1.7<sup>a</sup> | 3000<sup>a</sup> |
| $V_{mps}$; Volume of MPS (mL) | 1.56<sup>f</sup> | |
| $V_{lu}$; Volume of lungs (mL) | 0.5144<sup>f</sup> | 1170<sup>f</sup> |
| $V_k$; Volume of kidneys (mL) | 0.4956<sup>f</sup> | 280<sup>f</sup> |
| $V_o$; Volume of others (mL) | 13.2<sup>f</sup> | |
| Blood flow through lungs (mL/hr) | 667<sup>b</sup> | 330000<sup>c</sup> |
| Blood flow through MPS (mL/hr) | 113.4<sup>a</sup> | 87000<sup>a</sup> |
| Blood flow through kidneys (mL/hr) | 78<sup>a</sup> | 74400<sup>a</sup> |
| Blood flow through others (mL/hr) | 60<sup>b</sup> | 167400<sup>b</sup> |

<sup>a</sup>Davies and Morris et al., Pharm Res. (1993); <sup>b</sup>Tian et al. 2012 (30); <sup>c</sup>Ivanov et al., Bull Exp Biol Med. (2013); <sup>d</sup>De Ferraris et al., Médica panamericana. (1999); <sup>e</sup>Pankow et al., Health Phys. (1985). <sup>f</sup>Organ volumes were equal to their corresponding weights based on the assumption of organ density to be 1 g/mL.

### Interspecies scaling

To assess the translational potential of our mPBPK model for predicting SiNP kinetics at the whole-body level, we extrapolated the mouse model to humans. This was achieved by substituting the values of mouse parameters with human population averages from the literature (Table 2), or allometrically scaling the unknown parameters from mice to humans using established scaling factors. Rate constants were allometrically scaled based on established methods (33), using the following equation:

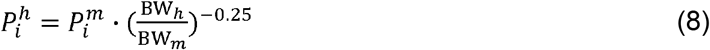

where, 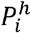 and 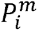 is the value of parameter *i* for humans and mice, respectively; *BW*_*h*_ and *BW*_*m*_ is the body weight of humans (assumed to be 70 kg) and mice (assumed to be 0.02 kg), respectively. The standardized allometric scaling exponent for rate constants is -0.25 (34). For dose and clearance calculations, an exponent of 0.75 is used, and for volume of distribution, an exponent of 1 is applied.

Note that the mouse physiological parameters for the model were obtained from the literature (35-39) and laboratory data (Table 2). Following allometric scaling, the human model parameters were recalibrated through least squares fitting using clinical data for C dots (Table 3).(30)

**Table 3.** mPBPK model parameter estimates from least squares regression.

| Parameter | Definition (Units) | Mice |  |  |  |  |  | Humans |
| --- | --- | --- | --- | --- | --- | --- | --- | --- |
|  |  | 8A | AR8 | MA | Meso | SA | Stöber | C dots |
| $f_{lu}$ | Unbound fraction of NPs in lungs (n/a) | 0.121 | 0.011 | 0.999 | 0.006 | 0.718 | 0.478 | 0.889 |
| $f_{mps}$ | Unbound fraction of NPs in MPS (n/a) | 0.073 | 0.003 | 0.497 | 0.005 | 0.0057 | 0.002 | 0.155 |
| $f_k$ | Unbound fraction of NPs in kidneys (n/a) | 0.907 | 0.596 | 0.926 | 0.513 | 0.747 | 0.541 | 0.078 |
| $f_o$ | Unbound fraction of NPs in others (n/a) | 0.541 | 0.029 | 0.508 | 0.065 | 0.083 | 0.019 | 0.964 |
| $Q_{mps}$ | Blood flow rate in MPS (mL/h) | 4.167 | 19.022 | 0.032 | 3.082 | 60.98 | 35.608 | 80854 |
| $Q_o$ | Blood flow rate in others (mL/h) | 9.559 | 43.492 | 6.962 | 1.649 | 76.07 | 84.858 | 165380 |
| $k_{e,mps}$ | Excretion rate constant in the MPS ($h^{-1}$) | 0.032 | 0.016 | 0.009 | 0.01 | 0.0025 | 0.003 | 0.452 |
| $k_{e,k}$ | Excretion rate constant in the kidneys ($h^{-1}$) | 27.342 | 21.679 | 32.578 | 3.357 | 12.18 | 15.443 | 2.67e-4 |

### Global sensitivity analysis

Global sensitivity analysis (GSA) was performed for the mouse model to quantify the influence of the eight fitted model parameters on the model’s output, specifically the area under the curve (AUC) for each compartment (from 0 to 500 hours). GSA was conducted using Latin hypercube sampling (LHS), a statistical method that generates a sample of plausible parameter values from a multidimensional parameter distribution.(40, 41) In total, 1,000 parameter combinations were generated over 10 replicates using LHS, with parameters perturbed between ±50% of their baseline values. Multivariate linear regression analysis (MLRA) was performed on each sample to generate sensitivity indices (SIs) for each model output, with the regression coefficients serving as the SIs.

The parameters were then ranked based on the SIs using one-way ANOVA and Tukey’s test across ten replicates. The ranking identified the most influential parameters by examining the SI distribution for each parameter. A higher SI indicates a greater influence of the corresponding parameter on the model’s AUC outputs. This global approach was chosen over local sensitivity analysis, which only perturbs one parameter at a time, as it accounts for potential interactions and interdependencies between parameters.

### Non-compartmental analysis

Non-compartmental analysis (NCA) was performed using PKanalix version 2021R2 (Antony, France, Lixoft SAS, 2021, http://lixoft.com/products/PKanalix/) software. The area under the curve (AUC) was calculated using the Linear Trapezoidal Linear method of PKanalix. The NCA was performed using plasma, MPS, lung, and kidney data. Reported data for mice and humans (in %ID/g units) was converted to concentration units for compatibility with PKanalix. The mean value for n=5 mice is used for NCA due to the unavailability of individual mouse data; therefore, there is no error or standard deviation to report.

## Results and Discussion

### Model Development

We developed a minimal PBPK (mPBPK) model to simulate the whole-body disposition kinetics of SiNPs with different physicochemical properties following I.V. administration in mice. The model was fitted to preclinical data for various SiNPs in mice (30) and clinical data for C dots in humans(31). Allometric scaling was then used to extrapolate the mouse model to humans. As shown by the PK profiles of various SiNPs in mice (Figure 2) and C dots in humans (Figure 3), the model accurately fits the disposition of these NPs across seven compartments in mice and six compartments in humans. Model parameter estimates obtained through non-linear least squares fitting for both mice and humans are provided in Table 3. The Pearson correlation coefficients for the different types of mesoporous SiNPs—Meso, MA, Stöber, SA, AR8, and 8A—are 0.99, 0.98, 0.98, 0.98, 0.98, 0.97, respectively. For the mean of the five human subjects, the Pearson correlation coefficient was 0.92, indicating the model’s strong agreement with the data. The experimental data consistently fell within the 95% confidence intervals derived from the model, underscoring its robustness and accuracy in predicting NP biodistribution in essential organs under varying physiological conditions and NP physicochemical properties. While this model adopts a minimalistic approach, it successfully captures the essential dynamics, making it a valuable tool for preliminary assessments and guiding more detailed experimental designs.

**Figure 2.**
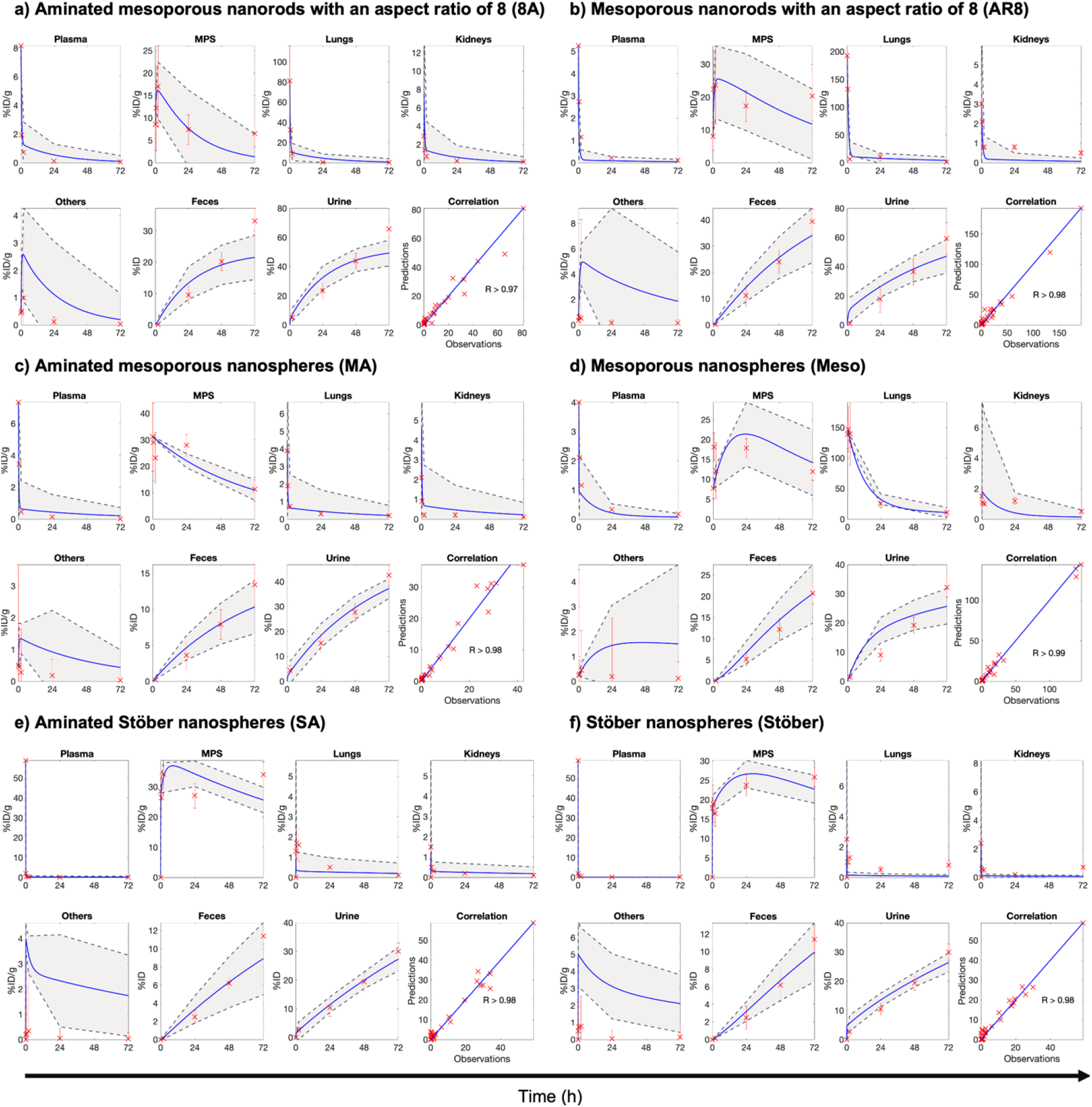
mPBPK model fits for various SiNPs in mice. The temporal kinetics of NP concentration (%ID/g) in plasma, MPS, lungs, kidneys, and ‘others’ compartment is shown for: **a)** mesoporous nanospheres (Meso), **b)** aminated mesoporous nanospheres (MA), **c)** Stöber nanospheres (Stöber), **d)** aminated Stöber nanospheres (SA), **e)** mesoporous nanorods with an aspect ratio of 8 (AR8), and **f)** aminated mesoporous nanorods with an aspect ratio of 8 (8A). Additionally, the cumulative mass kinetics (%ID) of NP excretion in urine and feces are shown for all NPs. Blue lines represent model fits, and red crosses with error bars represent the mean ± SD of in vivo data (n=5 mice). The gray shaded areas correspond to the 95% confidence interval of the model fits. Goodness of fit is assessed by Pearson correlation coefficient (R), displayed for each fit. A common arrow marks the time axis across all plots but note that this time axis does not apply to the correlation plots. In vivo data was obtained from Tian et al. 2012 (30).

**Figure 3.**
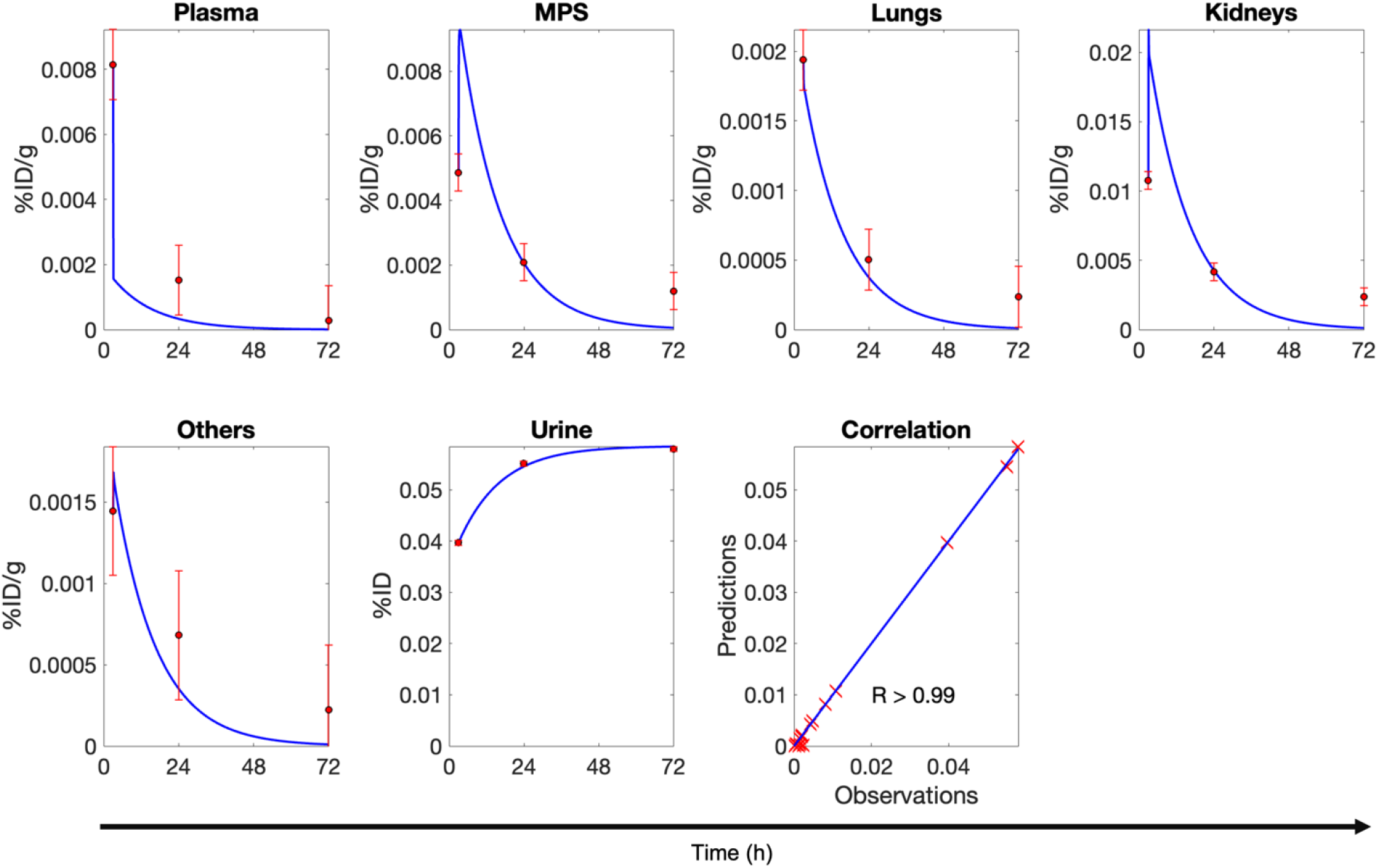
mPBPK model fits for C dots in humans. The temporal kinetics of NP concentration (%ID/g) in plasma, MPS, lungs, kidneys, and ‘others’ compartment is shown for C dots. Additionally, the cumulative mass kinetics (%ID) of NP excretion in urine is shown. Blue lines represent model fits, and red circles with error bars represent the mean ± SD of clinical data (n=5 subjects). Goodness of fit is assessed by Pearson correlation coefficient (R). A common arrow marks the time axis across all plots but note that this time axis does not apply to the correlation plots. Clinical data was obtained from Philips et al. 2014 (31).

**Figure 4.**
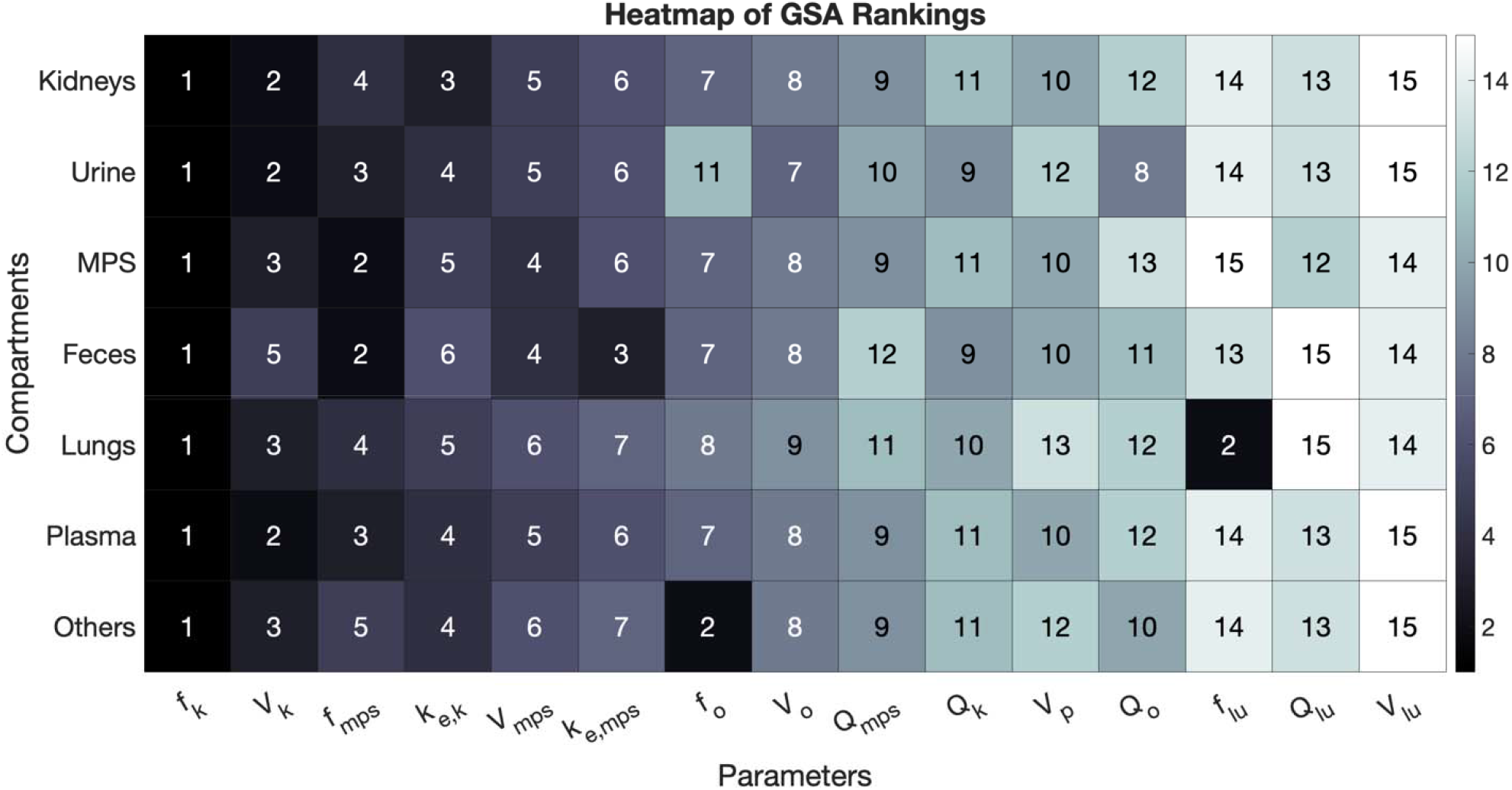
Global sensitivity analysis. Heat map of GSA-based rankings of various model parameters for their influence on the area under the curve (AUC) from 0–500 hours of NP concentration kinetics in the kidneys, plasma, lungs, MPS, and ‘others’ compartments in mice. For the urine and feces compartments, the cumulative mass at the final time point was the model output of interest. Darker colors and lower numbers represent higher sensitivity indices, indicating greater influence of the parameter on the model output.

### Global sensitivity analysis

The global sensitivity analysis (GSA) and exposure measurements (i.e., AUC_0–72 h_) from NCA provided detailed insights into how key model parameters influence the biodistribution of SiNPs across various compartments, including the plasma, kidneys, mononuclear phagocyte system (MPS), and lungs. The GSA highlighted the critical role of the fraction unbound in kidneys (*f*_*k*_) and fraction unbound in MPS (*f*_mps_), alongside the kidney elimination rate constant (*k*_*e,k*_) and MPS elimination rate constant (*k*_*e*,mps_). These parameters showed significant sensitivity across compartments and are strongly tied to the NP’s physicochemical properties, such as shape, porosity, surface charge, and coating. The violin plot of GSA-based rankings for the various model parameters in different model compartments of mice is shown in Supplementary Figure S1.

The model assumes that the unbound fraction (*f*_*k*_, *f*_mps_) represents the portion of NPs available for outflow via blood from a given organ, whereas the bound fraction (1− *f*_*k*_, 1− *f*_mps_) is subject to elimination through the respective elimination rate constants (*k*_*e,k*_ and *k*_*e*,mps_). NPs with higher unbound fractions tend to redistribute more readily through the bloodstream, reducing their retention in specific compartments, while lower unbound fractions lead to increased binding and retention. The elimination rate constants determine how quickly the bound fractions are cleared from the kidneys and MPS, impacting overall exposure.

Plasma exposure: The AUC values for plasma were generally lower than those for other compartments, but some variability was observed across NPs. MA and 8A exhibited the highest plasma exposure values (Figure 5), reflecting their high unbound fractions (Table 3). Stöber and AR8, on the other hand, showed lower plasma exposure, likely due to their low unbound fractions. This suggests that the plasma retention of these NPs is inversely related to their bound fractions.

**Figure 5.**
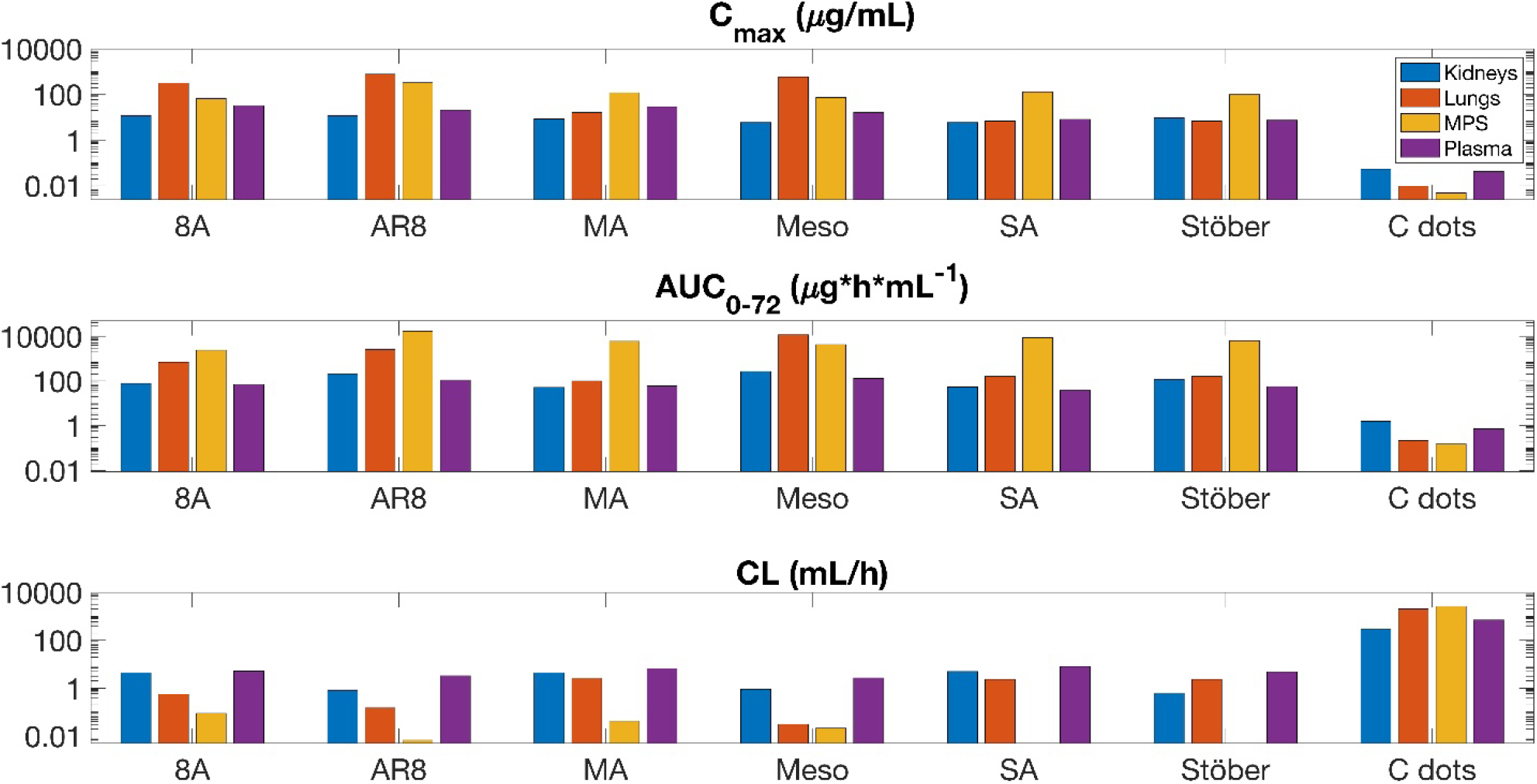
Non-compartmental analyses. Bar plot displaying results of NCA for each SiNP, including the following PK parameters: maximum concentration (C_max_), area under the concentration curve from time 0 to the final recorded timepoint (AUC_0-72_), and clearance (CL) in the plasma, lungs, MPS organs, and kidney compartments.

MPS Exposure: The MPS compartment demonstrated the highest AUC values for all NPs, indicating that the MPS plays a central role in NP retention. Among the NPs, SA and Stöber exhibited the greatest exposure in the MPS, with AR8 following closely behind. The slower elimination rates of these NPs from the MPS (*k*_*e*,mps_), particularly for SA, coupled with their lower unbound fractions (*f*_mps_), resulted in significant binding and prolonged retention.

In contrast, 8A had one of the lowest exposures in the MPS. This can be attributed to its higher unbound fraction and faster MPS elimination rate (*k*_*e*,mps_), which enabled it to accumulate less in the MPS. MA had slightly slower elimination, showed moderate MPS retention, reflecting the interplay between binding and elimination in determining MPS exposure.

Lung Exposure: The lung exposure values varied significantly across NPs, with AR8 and Meso, displaying the highest retention. AR8 and Meso benefit from their small unbound fractions in the lungs (*f*_lu_), which allows them to remain within lung tissues longer before redistribution. Additionally, AR8, being rod-shaped, may have unique interactions within the alveolar regions, slowing its clearance. Stöber and SA demonstrated the lowest lung retention, potentially due to their large unbound fractions (*f*_lu_) and more rapid processing through the MPS.

Kidney Exposure: The kidney exposure values showed lower AUCs than the MPS but still varied between NPs. Meso, 8A, and MA exhibited relatively higher kidney retention, driven by differences in their unbound fractions and kidney elimination rates. Meso demonstrated high retention in the kidneys due to its slower clearance, despite its small *f*_*k*_. The mesoporous structure of Meso likely contributes to its prolonged retention in the kidneys, as NPs with larger surface areas are often processed more slowly.

### Non-compartmental analysis

The results of the NCA for mice are shown in Table 4 and for humans in Table 5. It should be noted that clearance-related results of the NCA for the MPS compartment with Stöber NPs are unavailable due to an observed accumulation trend in MPS organs. As a result of the NCA, patterns in the PK of SiNPs were established based on the physical characteristics of SiNPs, including surface charge, size, and geometry. A visual representation in the form of a heat map of the SiNPs and the respective PK trend is shown in Figure 5.

**Table 4.** Non-compartmental analysis (NCA) of the disposition kinetics data for various SiNPs in mice. The maximum concentration (C_max_), time to maximum concentration (T_max_), area under the concentration curve from zero to infinity (AUC_0-∞_), area under the concentration curve from zero to seventy-two hours (AUC_0-72_), clearance (CL), volume of distribution at steady state (V_d,ss_), and half-life (t_1/2_) were calculated.

| NP Type | PK Parameters | Plasma | Lungs | MPS | Kidneys |
| --- | --- | --- | --- | --- | --- |
| **8A** | $C_{max}$ ($\mu\text{g/mL}$) | 33 | 320 | 66 | 12 |
| | $T_{max}$ (h) | 0 | 0 | 1.92 | 0 |
| | $AUC_{0-\infty}$ ($\text{h} \cdot \mu\text{g} \cdot \text{mL}^{-1}$) | 76 | 720 | 4540 | 91 |
| | $AUC_{0-72}$ ($\text{h} \cdot \mu\text{g} \cdot \text{mL}^{-1}$) | 71 | 710 | 2430 | 78 |
| | CL ($\text{mL/h}$) | 5.24 | 0.56 | 0.088 | 4.41 |
| | $V_{d,ss}$ (mL) | 123.42 | 7.63 | 7.38 | 136.27 |
| | $t_{1/2}$ (h) | 16.5 | 9.49 | 57.75 | 21.66 |
| **AR8** | $C_{max}$ ($\mu\text{g/mL}$) | 21 | 770 | 330 | 12 |
| | $T_{max}$ (h) | 0 | 0 | 0.42 | 0 |
| | $AUC_{0-\infty}$ ($\text{h} \cdot \mu\text{g} \cdot \text{mL}^{-1}$) | 120 | 2680 | 60050 | 490 |
| | $AUC_{0-72}$ ($\text{h} \cdot \mu\text{g} \cdot \text{mL}^{-1}$) | 110 | 2590 | 17200 | 210 |
| | CL ($\text{mL/h}$) | 3.3 | 0.15 | 0.0067 | 0.82 |
| | $V_{d,ss}$ (mL) | 84.43 | 2.72 | 1.31 | 114.23 |
| | $t_{1/2}$ (h) | 17.76 | 43.31 | 135.88 | 96.25 |
| **MA** | $C_{max}$ ($\mu\text{g/mL}$) | 29 | 16 | 120 | 8.4 |
| | $T_{max}$ (h) | 0 | 0 | 0 | 0 |
| | $AUC_{0-\infty}$ ($\text{h} \cdot \mu\text{g} \cdot \text{mL}^{-1}$) | 61 | 150 | 9600 | 90 |
| | $AUC_{0-72}$ ($\text{h} \cdot \mu\text{g} \cdot \text{mL}^{-1}$) | 60 | 100 | 6030 | 52 |
| | CL ($\text{mL/h}$) | 6.59 | 2.61 | 0.042 | 4.45 |
| | $V_{d,ss}$ (mL) | 104.88 | 159.23 | 3.4 | 418.7 |
| | $t_{1/2}$ (h) | 11 | 43.3 | 216.5 | 63 |
| **Meso** | $C_{max}$ ($\mu\text{g/mL}$) | 16 | 590 | 73 | 6 |
| | $T_{max}$ (h) | 0 | 0.42 | 0.42 | 0 |
| | $AUC_{0-\infty}$ ($\text{h} \cdot \mu\text{g} \cdot \text{mL}^{-1}$) | 150 | 12950 | 19040 | 440 |
| | $AUC_{0-72}$ ($\text{h} \cdot \mu\text{g} \cdot \text{mL}^{-1}$) | 130 | 11790 | 4290 | 270 |
| | CL ($\text{mL/h}$) | 2.66 | 0.031 | 0.021 | 0.91 |
| | $V_{d,ss}$ (mL) | 85.57 | 0.84 | 6.49 | 77.22 |
| | $t_{1/2}$ (h) | 22.35 | 18.72 | 216.56 | 57.75 |
| **SA** | $C_{max}$ ($\mu\text{g/mL}$) | 8.2 | 6.8 | 130 | 6 |
| | $T_{max}$ (h) | 0 | 0.42 | 1.92 | 0 |
| | $AUC_{0-\infty}$ ($\text{h} \cdot \mu\text{g} \cdot \text{mL}^{-1}$) | 49 | 170 | N/A | 81 |
| | $AUC_{0-72}$ ($\text{h} \cdot \mu\text{g} \cdot \text{mL}^{-1}$) | 38 | 160 | 8520 | 55 |
| | CL ($\text{mL/h}$) | 8.08 | 2.32 | N/A | 4.96 |
| | $V_{d,ss}$ (mL) | 344.76 | 58.61 | N/A | 320.02 |
| | $t_{1/2}$ (h) | 30.13 | 17.32 | N/A | 46.2 |
| **Stöber** | $C_{max}$ ($\mu\text{g/mL}$) | 7.5 | 6.8 | 100 | 9.6 |
| | $T_{max}$ (h) | 0 | 0.42 | 71.92 | 0 |
| | $AUC_{0-\infty}$ ($\text{h} \cdot \mu\text{g} \cdot \text{mL}^{-1}$) | 83 | 170 | N/A | 640 |
| | $AUC_{0-72}$ ($\text{h} \cdot \mu\text{g} \cdot \text{mL}^{-1}$) | 58 | 160 | 6450 | 120 |
| | CL ($\text{mL/h}$) | 4.84 | 2.32 | N/A | 0.62 |
| | $V_{d,ss}$ (mL) | 186.58 | 58.61 | N/A | 115.46 |
| | $t_{1/2}$ (h) | 26.65 | 17.32 | N/A | 128.33 |

**Table 5.**
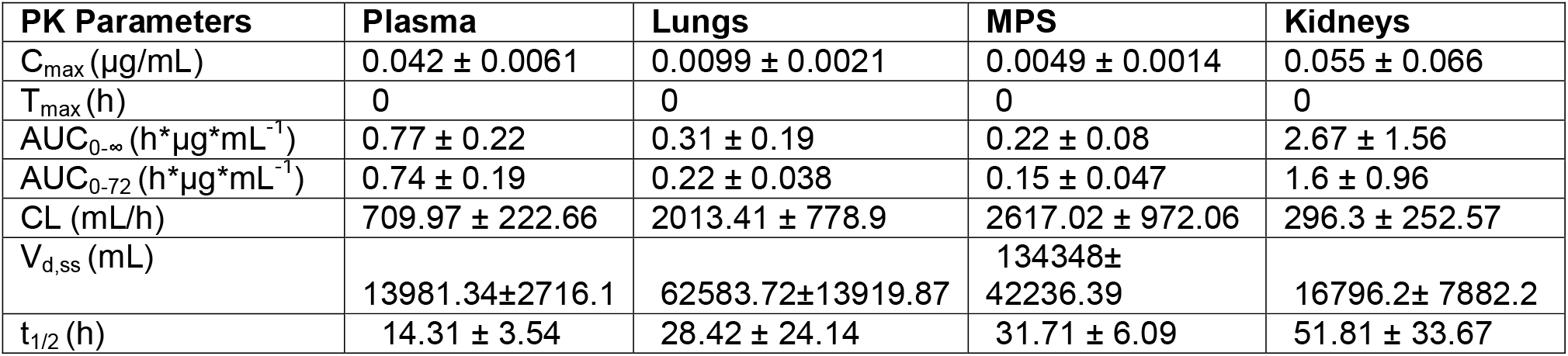
NCA of the disposition kinetics data for Cornell dots in humans. The maximum concentration (C_max_), time to maximum concentration (T_max_), area under the concentration curve from zero to infinity (AUC_0-∞_), area under the concentration curve from zero to seventy-two hours (AUC_0-72_), clearance (CL), volume of distribution at steady state (V_d,ss_), and half-life (t_1/2_) were evaluated.

The aminated SiNPS (8A, SA, and MA) tend to have the highest maximum concentration (C_max_) and area under the concentration curve from zero to infinity (AUC_0-_∞) and lowest clearance (CL) in the MPS and kidneys. Rod-shaped NPs tend to have a higher C_max_ in the lungs and plasma. MA and 8A SiNPs with the same highly positive zeta potential (20 to 40 mV), show the same magnitude C_max_, AUC_0-_∞, and CL in the plasma.

In this research, we collected data on the biodistribution of SiNPs from human and mouse subjects to develop a mPBPK model. This model integrates biodistribution and PK data to predict the behavior of diverse SiNP types in different biological systems. By comparing human and mouse data, we aimed to enhance the translation of SiNPs.

Since the process of extravasation involves the SiNPs moving from the bloodstream into the tissue, highly protein-bound SiNPs will have limited extravasation. Similarly, only unbound SiNPs can be recognized and internalized by macrophages. This will affect the clearance of the SiNPs as those with a higher fraction unbound will have higher uptake by the liver and spleen. For example, larger SiNPs have been established to reduce alveolar macrophage uptake.(44) Overall, capturing the unbound fraction via *in vitro* experiments would be necessary for a more meticulous model of NPs, including extravasation- and phagocytosis-dependent biodistribution and clearance.

The size and surface modification of SiNPs have been established to affect immune response.(45) (46) Small NPs are more likely to undergo phagocytosis and be cleared by the reticuloendothelial system (RES).(47) Smaller NPs have a higher surface area-to-volume ratio than larger particles. This increased surface area allows for more interactions with biological fluids, including plasma proteins and components of the immune system. Small NPs may pass through renal corpuscle cut-off (10 nm).(48) leading to increased, even rapid, clearance through the urine. (42) NPs with diameters exceeding this threshold are retained longer in the plasma due to decreased renal filtration. Interaction with renal tubules, which reabsorb water and nutrients, affects the excretion rates of NPs and their potential accumulation in the kidneys. Evidently, rapid clearance leads to reduced accumulation in organ tissues. This phenomenon is evidenced by the lowest AUC_0-_∞ and C_max_ and the quickest clearance of the C dots in all organs explored (plasma, lungs, MPS, kidney). This is supported by other NP studies that observed that other NPs have higher cellular uptake than smaller ones.(49, 50) The surface charge of the C dots administered in the source literature for this model was not reported. However, the surface conjugation of Cy5 dye contains negative sulfonate groups that will impart a negative charge.

Lung accumulation is observed with Meso and AR8 particles, two mesoporous SiNPs that are not amine-modified. AR8 and Meso have the highest AUC_0-_∞ and C_max_ and lowest CL in the lungs, likely due to their negative zeta potential (−40 to -30 mV). Amine modification reduces macrophage uptake in the lung. It reduces the risk of lung injury due to reduced induction of endosomal reactive oxygen species (ROS), leading to fewer ROS-dependent pathways for proinflammatory species induction.(44) It has been well documented that SiNPs of mesoporous nature preferentially accumulate in the lungs to do the lungs’ vascular nature, permeability, and capacity to retain substances and has led to an investigation of their use as drug delivery and imaging agents for lung-related ailments.(51-54)

Despite Stöber having a lower zeta potential than Meso and AR8 (−60 to -40 mV), lung accumulation is not observed. Notably, the nonporous version of Meso, Stöber, and Stöber’s aminated counterpart, SA, does not exhibit lung accumulation but instead accumulates in the MPS organs. The PK patterns of Stöber illustrate the complexity of PK when multiple physical properties are involved.

The mononuclear phagocyte system (MPS) plays a critical role in the plasma dynamics of NPs. Parameters such as k_e,MPS,_ and f_MPS_ directly influence NP levels through mechanisms of binding, uptake, and excretion. Upon entry into the bloodstream, NPs quickly bind to plasma proteins, forming a dynamic protein corona that modifies their biological identity and influences biodistribution and clearance. Coronas enriched with opsonins enhance phagocytic clearance by MPS cells. In organs like the liver and spleen, specialized endothelial structures and macrophages facilitate the capture and processing of NPs, leading to their excretion or degradation. A high k_e,MPS_ accelerates NP clearance from plasma, whereas a low k_e,MPS_ permits longer systemic circulation. The blood flow to MPS organs, while substantial, is typically not a limiting factor in NP clearance, assuming that perfusion is sufficient for effective NP delivery.

The mPBPK model was successfully created and verified with *in vivo* mouse and human biodistribution data. The Pearson correlation coefficient attests to a high level of agreement between the model predictions and observations.

To our knowledge, only one other SiNP PBPK model has been created that was specific to rats.(41) Other PBPK models for NPs have been created in MATLAB.(55-58) The current model does not include NP-specific parameters such as size, zeta potential, or geometry. We can establish patterns in biodistribution based on these parameters post-modeling. This study quantified the SiNPs in plasma and tissues using an indirect radioactive label-based method.

However, in our future studies, we would like to evaluate the PK of SiNPs and develop the PBPK model using PK data from a direct method of analysis using inductively coupled plasma mass spectrometry (ICP-MS).

## Conclusions

A robust PBPK model for SiNPs will significantly advance the field of nanomedicine by providing valuable insights into their PK in humans. In conclusion, this research successfully developed a PBPK model for SiNPs by integrating biodistribution data from both human and mouse subjects. The model highlighted key factors influencing NP behavior, including the role of protein binding in extravasation and phagocytosis and the impact of NP size and surface modifications on biodistribution and clearance. Future work will refine this model by incorporating direct ICP-MS analysis and exploring the delivery of drugs via SiNPs, enhancing its applicability and accuracy for predicting NP behavior in various biological systems. This research has the potential to impact the translational success of drug development, safety assessments, and regulatory evaluations of NP-based therapeutics, ultimately contributing to the safe and effective use of these innovative therapies.

## Funding Statement

This research was supported by NIH grant R03EB033576 (PI -VY, CO-I – PD and HG). The funders had no role in study design, data collection and analysis, publication decision, or manuscript preparation.

## Conflict of Interest Statement

The authors declare no conflicts of interest.

## Author Contributions

VY and PD conceived the study. PD developed the model and designed the analysis. MP, JC, MJP, PD, and VY performed the analysis. PD, HG, and VY interpreted the results. MP, JC, VY, and PD wrote the manuscript. MJP, and HG edited the manuscript.

## Data Availability Statement

The datasets are available from the corresponding author on reasonable request.

## Acknowledgements

PD and JC acknowledge Carmine Schiavone for helpful scientific discussions.

## Notes

**Conflict of Interest** Authors declare no conflict of interests.

### Competing Interest Statement

The authors have declared no competing interest.

### Funding Statement

This research was supported by NIH grant R03EB033576 (PI -VY). The funders had no role in study design, data collection and analysis, publication decision, or manuscript preparation.

### Author Declarations

https://www.ncbi.nlm.nih.gov/pmc/articles/PMC4426391/

## References

1. Liu Y, Mi Y, Zhao J, Feng SS. Multifunctional silica nanoparticles for targeted delivery of hydrophobic imaging and therapeutic agents. Int J Pharm. 2011;421(2):370–8.

2. Asad Mehmood HG, Samiya Yaqoob, Umar Farooq Gohar, Bashir Ahmad1. Mesoporous Silica Nanoparticles: A Review. Journal of Developing Drugs. 2017;6(2).

3. Benezra M, Penate-Medina O, Zanzonico PB, Schaer D, Ow H, Burns A, et al. Multimodal silica nanoparticles are effective cancer-targeted probes in a model of human melanoma. J Clin Invest. 2011;121(7):2768–80.

4. Huang X, Zhang F, Lee S, Swierczewska M, Kiesewetter DO, Lang L, et al. Long-term multimodal imaging of tumor draining sentinel lymph nodes using mesoporous silica-based nanoprobes. Biomaterials. 2012;33(17):4370–8.

5. Chen F, Hong H, Zhang Y, Valdovinos HF, Shi S, Kwon GS, et al. In vivo tumor targeting and image-guided drug delivery with antibody-conjugated, radiolabeled mesoporous silica nanoparticles. ACS Nano. 2013;7(10):9027–39.

6. Goel S, Chen F, Hong H, Valdovinos HF, Hernandez R, Shi S, et al. VEGF(1)(2)(1)-conjugated mesoporous silica nanoparticle: a tumor targeted drug delivery system. ACS Appl Mater Interfaces. 2014;6(23):21677–85.

7. Mohini Yadav VD, Swati Sharma, Nancy George,. Biogenic silica nanoparticles from agro-waste: Properties, mechanism of extraction and applications in environmental sustainability. Journal of Environmental Chemical Engineering. 2022;10(6).

8. Yang Y, Zhang M, Song H, Yu C. Silica-Based Nanoparticles for Biomedical Applications: From Nanocarriers to Biomodulators. Acc Chem Res. 2020;53(8):1545–56.

9. Kharlamov AN, Feinstein JA, Cramer JA, Boothroyd JA, Shishkina EV, Shur V. Plasmonic photothermal therapy of atherosclerosis with nanoparticles: long-term outcomes and safety in NANOM-FIM trial. Future Cardiol. 2017;13(4):345–63.

10. Kharlamov AN, Tyurnina AE, Veselova VS, Kovtun OP, Shur VY, Gabinsky JL. Silica– gold nanoparticles for atheroprotective management of plaques: results of the NANOM-FIM trial. Nanoscale. 2015;7(17):8003–15.

11. Huang Y, Li P, Zhao R, Zhao L, Liu J, Peng S, et al. Silica nanoparticles: Biomedical applications and toxicity. Biomed Pharmacother. 2022;151:113053.

12. Jeelani PG, Mulay P, Venkat R, Ramalingam C. Multifaceted Application of Silica Nanoparticles. A Review. Silicon. 2020;12:1337–54.

13. Hoshyar N, Gray S, Han H, Bao G. The effect of nanoparticle size on in vivo pharmacokinetics and cellular interaction. Nanomedicine (Lond). 2016;11(6):673–92.

14. Ridolfo R, Tavakoli S, Junnuthula V, Williams DS, Urtti A, van Hest JCM. Exploring the Impact of Morphology on the Properties of Biodegradable Nanoparticles and Their Diffusion in Complex Biological Medium. Biomacromolecules. 2021;22(1):126–33.

15. Xuan Y, Zhang W, Zhu X, Zhang S. An updated overview of some factors that influence the biological effects of nanoparticles. Front Bioeng Biotechnol. 2023;11:1254861.

16. Li M, Al-Jamal KT, Kostarelos K, Reineke J. Physiologically based pharmacokinetic modeling of nanoparticles. ACS Nano. 2010;4(11):6303–17.

17. Yuan D, He H, Wu Y, Fan J, Cao Y. Physiologically Based Pharmacokinetic Modeling of Nanoparticles. J Pharm Sci. 2019;108(1):58–72.

18. Kumar M, Kulkarni P, Liu S, Chemuturi N, Shah DK. Nanoparticle biodistribution coefficients: A quantitative approach for understanding the tissue distribution of nanoparticles. Adv Drug Deliv Rev. 2023;194:114708.

19. Li M, Zou P, Tyner K, Lee S. Physiologically Based Pharmacokinetic (PBPK) Modeling of Pharmaceutical Nanoparticles. AAPS J. 2017;19(1):26–42.

20. Chou W-C, Cheng Y-H, Riviere JE, Monteiro-Riviere NA, Kreyling WG, Lin Z. Development of a multi-route physiologically based pharmacokinetic (PBPK) model for nanomaterials: a comparison between a traditional versus a new route-specific approach using gold nanoparticles in rats. Particle and Fibre Toxicology. 2022;19(1).

21. Gonzalez-Valdivieso J, Girotti A, Schneider J, Arias FJ. Advanced nanomedicine and cancer: Challenges and opportunities in clinical translation. Int J Pharm. 2021;599:120438.

22. Metselaar JM, Lammers T. Challenges in nanomedicine clinical translation. Drug Deliv Transl Res. 2020;10(3):721–5.

23. Satalkar P, Elger BS, Hunziker P, Shaw D. Challenges of clinical translation in nanomedicine: A qualitative study. Nanomedicine. 2016;12(4):893–900.

24. Richfield O, Piotrowski-Daspit AS, Shin K, Saltzman WM. Rational nanoparticle design: Optimization using insights from experiments and mathematical models. J Control Release. 2023;360:772–83.

25. Krumpholz L, Polak S, Wisniowska B. Physiologically-based pharmacokinetic model of in vitro porcine ear skin permeation for drug delivery research. J Appl Toxicol. 2024.

26. Chang M, Chen Y, Ogasawara K, Schmidt BJ, Gaohua L. Advancements in physiologically based pharmacokinetic modeling for fedratinib: updating dose guidance in the presence of a dual inhibitor of CYP3A4 and CYP2C19. Cancer Chemother Pharmacol. 2024.

27. Rowland M, Peck C, Tucker G. Physiologically-based pharmacokinetics in drug development and regulatory science. Annu Rev Pharmacol Toxicol. 2011;51:45–73.

28. Miller NA, Reddy MB, Heikkinen AT, Lukacova V, Parrott N. Physiologically Based Pharmacokinetic Modelling for First-In-Human Predictions: An Updated Model Building Strategy Illustrated with Challenging Industry Case Studies. Clin Pharmacokinet. 2019;58(6):727–46.

29. Yu T, Malugin A, Ghandehari H. Impact of silica nanoparticle design on cellular toxicity and hemolytic activity. ACS Nano. 2011;5(7):5717–28.

30. Yu T, Hubbard D, Ray A, Ghandehari H. In vivo biodistribution and pharmacokinetics of silica nanoparticles as a function of geometry, porosity, and surface characteristics. J Control Release. 2012;163(1):46–54.

31. Phillips E, Penate-Medina O, Zanzonico PB, Carvajal RD, Mohan P, Ye Y, et al. Clinical translation of an ultrasmall inorganic optical-PET imaging nanoparticle probe. Science Translational Medicine. 2014;6(260):260ra149–260ra1.

32. Chen F, Ma K, Benezra M, Zhang L, Cheal SM, Phillips E, et al. Cancer-Targeting Ultrasmall Silica Nanoparticles for Clinical Translation: Physicochemical Structure and Biological Property Correlations. Chem Mater. 2017;29(20):8766–79.

33. Dogra P, Ramírez JR, Butner JD, Peláez MJ, Chung C, Hooda-Nehra A, et al. Translational Modeling Identifies Synergy between Nanoparticle-Delivered miRNA-22 and Standard-of-Care Drugs in Triple-Negative Breast Cancer. Pharmaceutical Research. 2022;39(3):511–28.

34. Lindauer A, Valiathan C, Mehta K, Sriram V, De Greef R, Elassaiss-Schaap J, et al. Translational Pharmacokinetic/Pharmacodynamic Modeling of Tumor Growth Inhibition Supports Dose-Range Selection of the Anti-PD-1 Antibody Pembrolizumab. CPT: Pharmacometrics & Systems Pharmacology. 2017;6(1):11–20.

35. De Ferraris MEG. Histología y embriología bucodental. Médica panamericana. 1999.

36. B.K.B. Berkovitz Grh, B.J. Moxham. A colour atlas & text of oral anatomy : histology and embryology. 2 ed: Wolfe; 1992.

37. Davies B, Morris T. Physiological parameters in laboratory animals and humans. Pharm Res. 1993;10(7):1093–5.

38. Pankow BG, Michalak J, McGee MK. Adult human thyroid weight. Health Phys. 1985;49(6):1097–103.

39. Ivanov KP. New data on the process of circulation and blood oxygenation in the lungs under physiological conditions. Bull Exp Biol Med. 2013;154(4):411–4.

40. Zhang S, Zhang E, Ho H. Extrapolation for a pharmacokinetic model for acetaminophen from adults to neonates: A Latin Hypercube Sampling analysis. Drug Metab Pharmacokinet. 2020;35(3):329–33.

41. Dogra P, Butner JD, Ruiz Ramirez J, Chuang YL, Noureddine A, Jeffrey Brinker C, et al. A mathematical model to predict nanomedicine pharmacokinetics and tumor delivery. Comput Struct Biotechnol J. 2020;18:518–31.

42. Adhipandito CF, Cheung S-H, Lin Y-H, Wu S-H. Atypical Renal Clearance of Nanoparticles Larger Than the Kidney Filtration Threshold. International Journal of Molecular Sciences. 2021;22(20):11182.

43. Dogra P, Adolphi NL, Wang Z, Lin YS, Butler KS, Durfee PN, et al. Establishing the effects of mesoporous silica nanoparticle properties on in vivo disposition using imaging-based pharmacokinetics. Nat Commun. 2018;9(1):4551.

44. Inoue M, Sakamoto K, Suzuki A, Nakai S, Ando A, Shiraki Y, et al. Size and surface modification of silica nanoparticles affect the severity of lung toxicity by modulating endosomal ROS generation in macrophages. Particle and Fibre Toxicology. 2021;18(1).

45. Murugadoss S, Lison D, Godderis L, Van Den Brule S, Mast J, Brassinne F, et al. Toxicology of silica nanoparticles: an update. Archives of Toxicology. 2017;91(9):2967–3010.

46. Alessandrini, Aguilar Pimentel JA, Landsiedel R, Wohlleben W, Mempel M, Marzaioli V, et al. Surface modifications of silica nanoparticles are crucial for their inert versus proinflammatory and immunomodulatory properties. International Journal of Nanomedicine. 2014:2815.

47. Yu M, Zheng J. Clearance Pathways and Tumor Targeting of Imaging Nanoparticles. ACS Nano. 2015;9(7):6655–74.

48. Choi CH, Zuckerman JE, Webster P, Davis ME. Targeting kidney mesangium by nanoparticles of defined size. Proc Natl Acad Sci U S A. 2011;108(16):6656–61.

49. Limbach LK, Li Y, Grass RN, Brunner TJ, Hintermann MA, Muller M, et al. Oxide nanoparticle uptake in human lung fibroblasts: effects of particle size, agglomeration, and diffusion at low concentrations. Environ Sci Technol. 2005;39(23):9370–6.

50. Ge Y, Zhang Y, Xia J, Ma M, He S, Nie F, et al. Effect of surface charge and agglomerate degree of magnetic iron oxide nanoparticles on KB cellular uptake in vitro. Colloids Surf B Biointerfaces. 2009;73(2):294–301.

51. Cheng W, Liang C, Xu L, Liu G, Gao N, Tao W, et al. TPGS-Functionalized Polydopamine-Modified Mesoporous Silica as Drug Nanocarriers for Enhanced Lung Cancer Chemotherapy against Multidrug Resistance. Small. 2017;13(29):1700623.

52. Min S, Tao W, Miao Y, Li Y, Wu T, He X, et al. Dual Delivery of Tetramethylpyrazine and miR-194-5p Using Soft Mesoporous Organosilica Nanoparticles for Acute Lung Injury Therapy. Int J Nanomedicine. 2023;18:6469–86.

53. Campos Pacheco JE, Yalovenko T, Riaz A, Kotov N, Davids C, Persson A, et al. Inhalable porous particles as dual micro-nano carriers demonstrating efficient lung drug delivery for treatment of tuberculosis. Journal of Controlled Release. 2024;369:231–50.

54. Garcia-Fernandez A, Sancho M, Bisbal V, Amoros P, Marcos MD, Orzaez M, et al. Targeted-lung delivery of dexamethasone using gated mesoporous silica nanoparticles. A new therapeutic approach for acute lung injury treatment. J Control Release. 2021;337:14–26.

55. Kagan L, Gershkovich P, Wasan KM, Mager DE. Dual physiologically based pharmacokinetic model of liposomal and nonliposomal amphotericin B disposition. Pharm Res. 2014;31(1):35–45.

56. Dong D, Wang X, Wang H, Zhang X, Wang Y, Wu B. Elucidating the in vivo fate of nanocrystals using a physiologically based pharmacokinetic model: a case study with the anticancer agent SNX-2112. Int J Nanomedicine. 2015;10:2521–35.

57. Aborig M, Malik PRV, Nambiar S, Chelle P, Darko J, Mutsaers A, et al. Biodistribution and Physiologically-Based Pharmacokinetic Modeling of Gold Nanoparticles in Mice with Interspecies Extrapolation. Pharmaceutics. 2019;11(4):179.

58. Howell BA, Chauhan A. A physiologically based pharmacokinetic (PBPK) model for predicting the efficacy of drug overdose treatment with liposomes in man. J Pharm Sci. 2010;99(8):3601–19.

