## Supplementary Figure S1 for "A Minimal PBPK Model Describes the Differential Disposition of Silica Nanoparticles In Vivo"

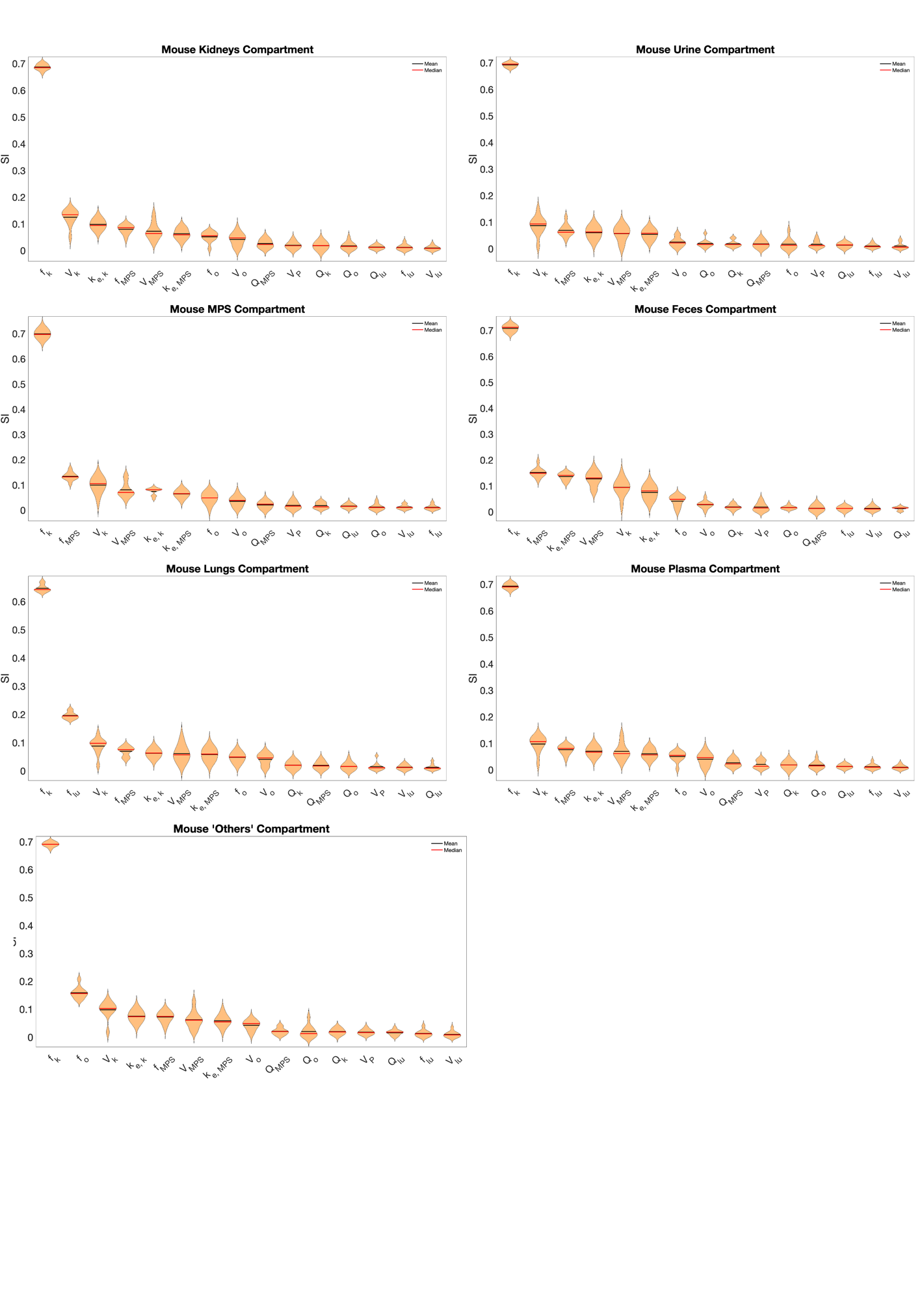
**Figure S1. Violin plot of GSA-based rankings for the various model parameters in different model compartments of mice.**
